# Online grocery shopping during the COVID-19 pandemic - perspectives of adolescents and young adults

**DOI:** 10.1101/2022.06.30.22277101

**Authors:** Eric J Brandt, Nicole Hadler, Ione Locher, Charlie T Hoffs, Marika E Waselewski, Tammy Chang

**Affiliations:** Institute for Healthcare Policy and Innovation, University of Michigan, Ann Arbor, MI; Division of Cardiovascular Medicine, Department of Internal Medicine, University of Michigan, Ann Arbor, MI; University of Michigan Medical School, Ann Arbor, MI; Stanford University, Stanford, CA; Department of Family Medicine, University of Michigan, Ann Arbor, MI

**Keywords:** youth, SNAP, qualitative, grocery

## Abstract

**Objective:** To assess perspectives of online grocery shopping during the COVID-19 pandemic among youth, rural residents, and Supplemental Nutrition Assistance Program (SNAP) participants.

**Design:** Open-ended text message survey data. Survey questions assessed rates of use and perspectives of online grocery shopping among youth and their families during the COVID-19 pandemic. Qualitative analysis of survey data from 875 participants (response rate=76.4%) to identify themes in experience with multivariable logistic regression to test associations between online grocery shopping (pickup, delivery, or either) with rurality and SNAP participation.

**Setting:** United States

**Participants:** Nationwide text-messaging poll of youth (14-24 years-old) recruited to meet benchmarks based on the American Community Survey.

**Results:** During the pandemic online grocery shopping was used frequently (46.7%). Safety and convenience were the primary reasons for preferring a shopping mode (in-person or online). Most online shoppers had positive experiences (59.4%), primarily due to convenience; negative experiences (28.3%) were from inaccuracies, inconveniences of the process, and delivery costs. Rural and suburban residence was associated with higher pickup (OR 2.02 and 1.51, respectively, p=.03) and lower delivery use (OR 0.33 and 0.72, respectively, p=.003) compared to urban residence. SNAP participation was not associated with any type of online grocery shopping.

**Conclusions:** Online grocery shopping is common among youth and their families regardless of rurality or SNAP participation, but there are several youth-identified areas for improvement.

**GRAPHICAL ABSTRACT:** 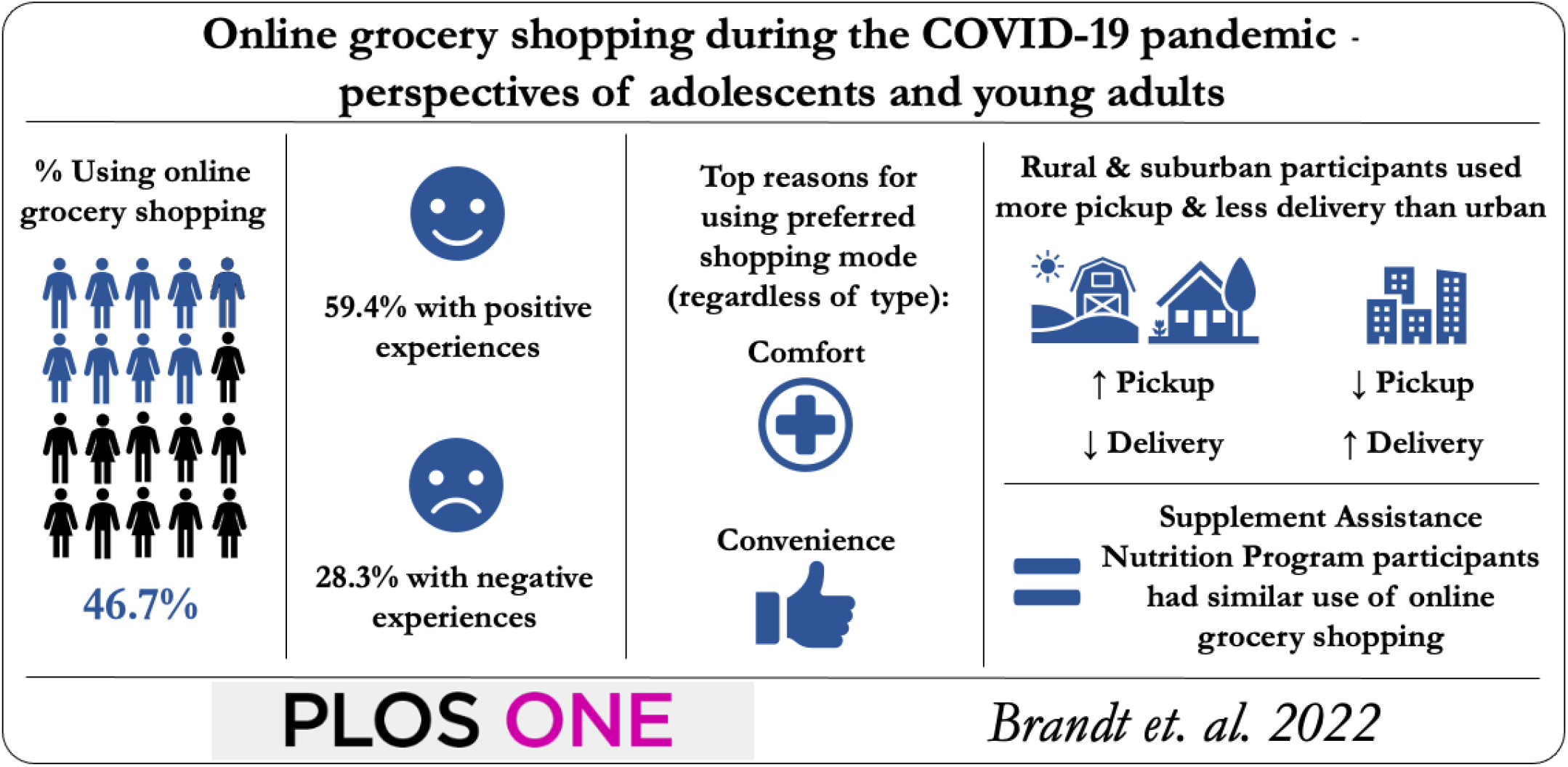

## Introduction

Throughout the pandemic, the Centers for Disease Control and Prevention has recommended online grocery shopping to mitigate the spread of COVID-19.^1^ Online grocery shopping grew tremendously, largely from concerns about contracting COVID-19 from in-person shopping.^2-4^ In addition, online grocery shopping access has expanded for Supplemental Nutrition Assistance Program (SNAP) participants, although rural residents may have lower access to grocery delivery.^3^ With diet as the top contributor to premature death in the US, online grocery shopping provides an opportunity to maximize its equitable use for healthy food access.^5,6^ However, there are several perceived disadvantages to its use, including cost, geography, browsing limitations, and trust, which may be heightened among vulnerable populations.^7-10^

Before the pandemic, families with children were the highest users of online grocery shopping.^11^ There is a paucity of literature about online grocery shopping among adolescent and young adults, especially among vulnerable populations and during the pandemic. We sought to assess the rates of use and perspectives of online grocery shopping among youth and their families during the COVID-19 pandemic. We hypothesized that due to barriers in online grocery shopping, its use would be lower among those in rural settings and among those who participate in SNAP.

## Methods

Respondents are part of the MyVoice cohort, an ongoing nationwide text-message poll seeking US youth opinions on health and policy issues.^12^ Participants are 14 to 24 years old and recruited on a rolling basis to meet national demographic benchmarks (age, sex, race/ethnicity, and region) based on weighted samples of the American Community Survey and receive $1 for each week they answer survey questions. Survey questions were developed in collaboration with experts in survey design, adolescent health, and qualitative and mixed methods research, as well as youth. On November 13, 2020, we posed three open-ended questions to this cohort via text message and respondents had 1 week to answer:

1. “During the pandemic, how have you or your family been getting groceries (in-person, pickup, delivery)? Why?”
2. “If you or your family have shopped for groceries online during the pandemic, was it pickup or delivery? How did it go?”
3. “During the pandemic, have you or your family changed the types of food you are buying (more/less snacks, fruits, vegetables, etc.)? Tell us about it.”

Respondents also reported the rurality of their community (rural, suburban, or urban) and household SNAP participation. Demographic information was obtained at enrollment. Themes and codes were identified and discussed to generate a codebook for each question. Responses were then analyzed and coded independently by two investigators using a modified grounded theory approach. Differences were discussed to reach agreement. Summary statistics of demographics and code frequencies were calculated. We used multivariable logistic regression to test associations between online grocery shopping (pickup, delivery, or either) with rurality and SNAP participation. P-values were 2-tailed and statistical significance was set at <0.05. Analyses were controlled for age, sex, race, and ethnicity, and completed using Stata 16 (StataCorp, LLC.). This study was approved by the University of Michigan Institutional Review Board, including a waiver of parental consent for minor participants.

## Results

Among 875 respondents (response rate=76.4%), median age was 18 years, 47.0% were female, 65.5% were white, 12.8% were Hispanic, 12.9% were from rural areas, and 12.8% of households participated in SNAP (Table 1).

**Table 1:**
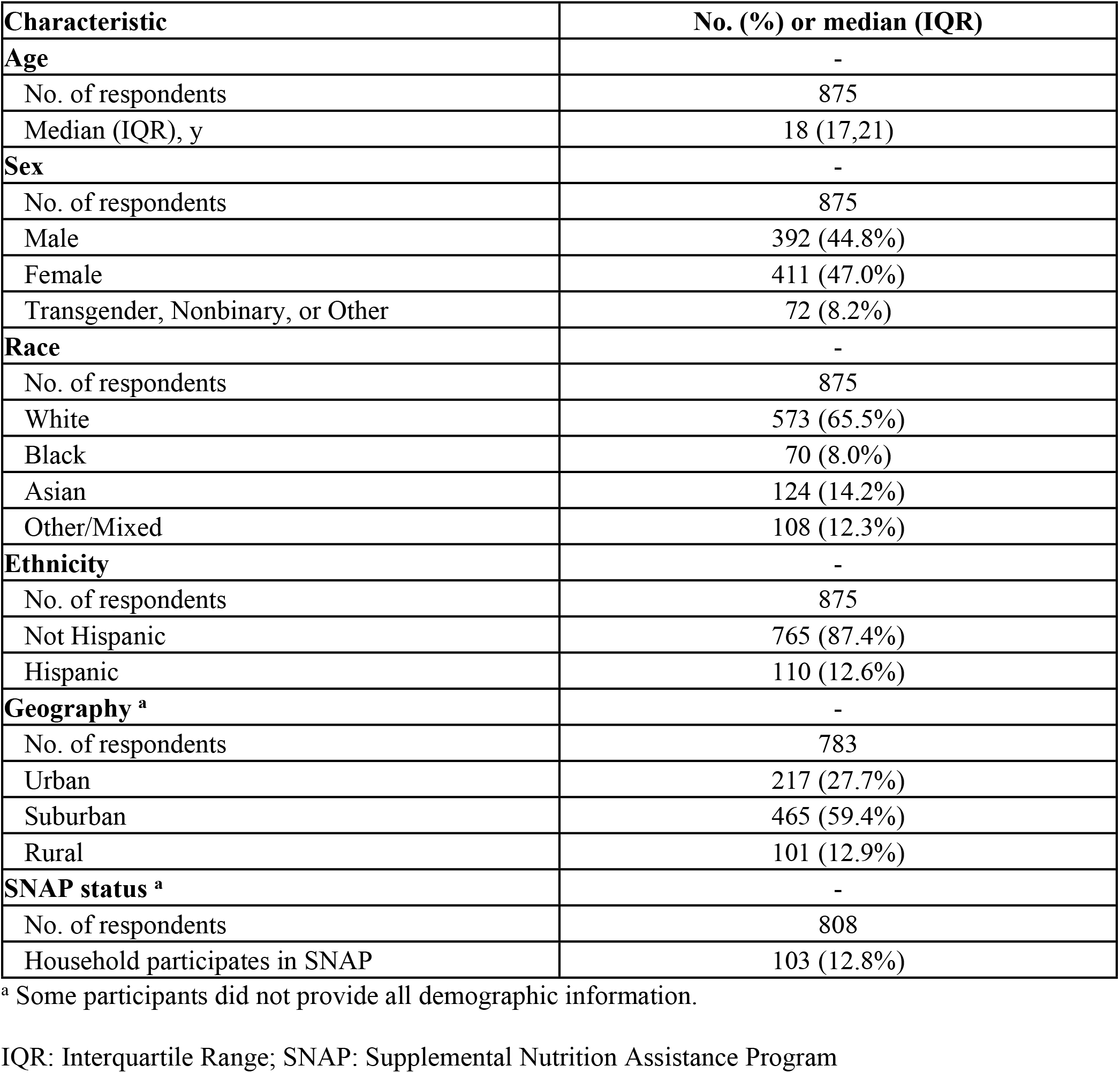
Demographic Characteristics of the Study Sample

| **Characteristic** | **No. (%) or median (IQR)** |
| --- | --- |
| **Age** | - |
| No. of respondents | 875 |
| Median (IQR), y | 18 (17,21) |
| **Sex** | - |
| No. of respondents | 875 |
| Male | 392 (44.8%) |
| Female | 411 (47.0%) |
| Transgender, Nonbinary, or Other | 72 (8.2%) |
| **Race** | - |
| No. of respondents | 875 |
| White | 573 (65.5%) |
| Black | 70 (8.0%) |
| Asian | 124 (14.2%) |
| Other/Mixed | 108 (12.3%) |
| **Ethnicity** | - |
| No. of respondents | 875 |
| Not Hispanic | 765 (87.4%) |
| Hispanic | 110 (12.6%) |
| **Geography <sup>a</sup>** | - |
| No. of respondents | 783 |
| Urban | 217 (27.7%) |
| Suburban | 465 (59.4%) |
| Rural | 101 (12.9%) |
| **SNAP status <sup>a</sup>** | - |
| No. of respondents | 808 |
| Household participates in SNAP | 103 (12.8%) |

<sup>a</sup> Some participants did not provide all demographic information.
IQR: Interquartile Range; SNAP: Supplemental Nutrition Assistance Program

Grocery shopping modes during the pandemic were not mutually exclusive, and 46.7% noted using online grocery shopping at least once (27.2% pickup; 26.3% delivery). Reasons for using online grocery shopping included safety (37.1%), convenience (20.1%), and transportation limitations (3.4%) (Table 2). Reasons for in-person shopping included being perceived as safe (23.4%), more convenient (20.3%), picking one’s own food (11.8%), and affordability (11.6%). Individuals also described in-person shopping as providing a sense of normalcy (5.3%), online grocery shopping being unavailable (4.6%), or not fearing COVID (4.4%).

**Table 2:**
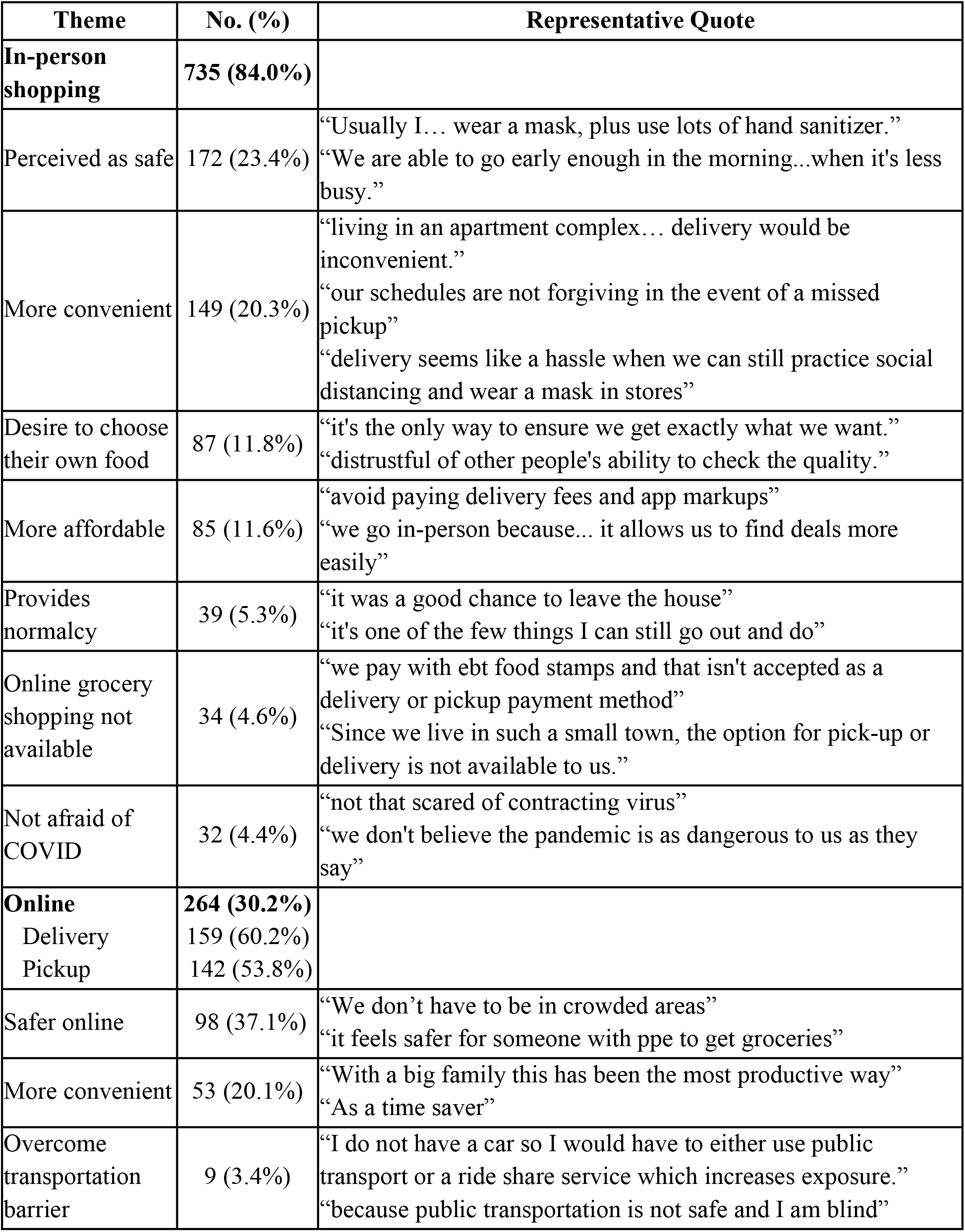
Reported Use of In-person and Online Grocery Shopping and Reasons for Use

| Theme | No. (%) | Representative Quote |
| --- | --- | --- |
| **In-person shopping** | **735 (84.0%)** |  |
| Perceived as safe | 172 (23.4%) | “Usually I... wear a mask, plus use lots of hand sanitizer.”<br>“We are able to go early enough in the morning...when it's less busy.” |
| More convenient | 149 (20.3%) | “living in an apartment complex... delivery would be inconvenient.”<br>“our schedules are not forgiving in the event of a missed pickup”<br>“delivery seems like a hassle when we can still practice social distancing and wear a mask in stores” |
| Desire to choose their own food | 87 (11.8%) | “it's the only way to ensure we get exactly what we want.”<br>“distrustful of other people's ability to check the quality.” |
| More affordable | 85 (11.6%) | “avoid paying delivery fees and app markups”<br>“we go in-person because... it allows us to find deals more easily” |
| Provides normalcy | 39 (5.3%) | “it was a good chance to leave the house”<br>“it's one of the few things I can still go out and do” |
| Online grocery shopping not available | 34 (4.6%) | “we pay with ebt food stamps and that isn't accepted as a delivery or pickup payment method”<br>“Since we live in such a small town, the option for pick-up or delivery is not available to us.” |
| Not afraid of COVID | 32 (4.4%) | “not that scared of contracting virus”<br>“we don't believe the pandemic is as dangerous to us as they say” |
| **Online** | **264 (30.2%)** |  |
| Delivery | 159 (60.2%) |  |
| Pickup | 142 (53.8%) |  |
| Safer online | 98 (37.1%) | “We don't have to be in crowded areas”<br>“it feels safer for someone with ppe to get groceries” |
| More convenient | 53 (20.1%) | “With a big family this has been the most productive way”<br>“As a time saver” |
| Overcome transportation barrier | 9 (3.4%) | “I do not have a car so I would have to either use public transport or a ride share service which increases exposure.”<br>“because public transportation is not safe and I am blind” |

Experiences with online grocery shopping were largely positive (59.4%), most commonly for convenience (47.8% of positive responses; “saved us a ton of time” or “…was super easy. Just a click and delivery”). Negative experiences (28.3%) were related to inaccuracies (57.8% of negative responses; “missing items” or “substitutes we got were not useful”), inconveniences (28.4%; “pickup was slow” or “difficulties in tech”), cost (11.9%; “way too expensive” or “you can’t use coupons”), and product quality problems (9.1%; “they give you rotten produce”).

Approximately half (50.4%) of respondents reported changing food purchasing habits during the pandemic. The most common report were of more healthier (32.5%) than less healthy purchases (11.4%), more increased (26.5%) than decreased snack foods (8.5%), and more purchases of shelf-stable foods (18.2%).

Rurality of the respondent’s community predicted using pickup (rural OR=2.02; suburban OR=1.51 versus urban, p=.03) and delivery (rural OR=0.33; suburban OR=0.72 versus urban, p=.003), but not online grocery shopping overall (pickup and delivery combined; Table 3). Household SNAP participation did not associate with use of online grocery shopping.

**Table 3:** Association of Shopping Mode with Rurality and SNAP Participation in Multivariable Logistic Regression Models

| A: Rurality |  |  |  |  |  |  |
| --- | --- | --- | --- | --- | --- | --- |
|  | Online Grocery Shopping<br>OR (95% CI) | p-value | Pickup<br>OR (95% CI) | p-value | Delivery<br>OR (95% CI) | p-value |
| Rurality |  | 0.48 |  | **0.03\*** |  | **0.003\*\*** |
| Urban (ref.) |  |  |  |  |  |  |
| Suburban | 0.86 (0.62, 1.20) |  | 1.51 (1.01, 2.26) |  | 0.72 (0.50, 1.04) |  |
| Rural | 0.75 (0.63, 1.57) |  | 2.02 (1.17, 3.50) |  | 0.33 (0.17, 0.62) |  |
| Age >18 | 1.04 (0.76, 1.41) | 0.83 | 0.94 (0.66, 1.33) | 0.72 | 1.13 (0.79, 1.62) | 0.49 |
| Sex |  | 0.83 |  | 0.22 |  | 0.77 |
| Male (ref.) |  |  |  |  |  |  |
| Female | 0.91 (0.67, 1.23) |  | 0.78 (0.55, 1.11) |  | 1.07 (0.75, 1.51) |  |
| Nonbinary | 0.94 (0.54, 1.65) |  | 0.63 (0.32, 1.23) |  | 1.24 (0.67, 2.29) |  |
| Race |  | 0.15 |  | **0.02\*** |  | 0.42 |
| White (ref.) |  |  |  |  |  |  |
| Black | 1.15 (0.67, 2.00) |  | 1.21 (0.66, 2.22) |  | 0.84 (0.45, 1.55) |  |
| Asian | 0.63 (0.41, 0.96) |  | 0.43 (0.24, 0.75) |  | 0.72 (0.44, 1.17) |  |
| Other | 0.99 (0.63, 1.57) |  | 1.02 (0.61, 1.71) |  | 0.73 (0.43, 1.26) |  |
| Hispanic | 0.90 (0.57, 1.43) | 0.67 | 0.75 (0.44, 1.29) | 0.30 | 1.16 (0.69, 1.94) | 0.57 |
| B: SNAP Participation |  |  |  |  |  |  |
|  | Online Grocery Shopping<br>OR (95% CI) | p-value | Pickup<br>OR (95% CI) | p-value | Delivery<br>OR (95% CI) | p-value |
| Household Participates in SNAP | 0.72 (0.47, 1.12) | 0.15 | 0.94 (0.57, 1.55) | 0.80 | 0.65 (0.39, 2.15) | 0.11 |
| Age >18 | 1.05 (0.78, 1.42) | 0.75 | 0.83 (0.59, 1.17) | 0.29 | 1.30 (0.92, 1.83) | 0.14 |
| Sex |  | 0.95 |  | 0.33 |  | 0.84 |
| Male (ref.) |  |  |  |  |  |  |
| Female | 0.95 (0.70, 1.29) |  | 0.83 (0.59, 1.17) |  | 1.08 (0.77, 1.52) |  |
| Nonbinary | 0.99 (0.57, 1.72) |  | 0.65 (0.34, 1.26) |  | 1.16 (0.63, 2.14) |  |
| Race |  | 0.60 |  | **0.03\*** |  | 0.64 |
| White (ref.) |  |  |  |  |  |  |
| Black | 1.32 (0.76, 2.30) |  | 1.11 (0.60, 2.04) |  | 1.07 (0.57, 1.99) |  |
| Asian | 0.67 (0.44, 1.02) |  | 0.45 (0.26, 0.77) |  | 0.81 (0.50, 1.31) |  |
| Other | 1.00 (0.64, 1.57) |  | 0.99 (0.60, 1.65) |  | 0.77 (0.46, 1.31) |  |
| Hispanic | 0.96 (0.61, 1.50) | 0.85 | 0.70 (0.41, 1.19) | 0.19 | 1.31 (0.80, 2.15) | 0.29 |

Boldface indicates statistical significance (\*p<0.05, \*\*p<0.01).
SNAP, Supplemental Nutrition Assistance Program

## Discussion

Youth and their families commonly used online grocery shopping during the COVID-19 pandemic. The majority of respondents stated that safety and convenience were the primary reasons for their choice to shop for groceries either in-person or online. Despite a majority of individuals having positive experiences with online grocery shopping there were several areas for improvement noted. Respondents’ rurality was associated with the type of online grocery shopping utilized (pickup or delivery) but not any use of online grocery shopping. Household SNAP participation was not associated with use of online grocery shopping.

Youth responses identified areas for improvement, which included increasing delivery availability, improving systems to prevent inaccuracies, increasing food options, guaranteeing food quality, and achieving price parity to in-store prices. Several of the youth-identified pitfalls align with previously identified opportunities to improve online grocery shopping, including increasing delivery availability, improving systems to prevent inaccuracies, increasing food options, guaranteeing food quality, and achieving price parity to in-store prices.^13^ Focusing on these modifications may improve comfort in use of online grocery shopping both as a convenience tool and to help mitigate spread of COVID-19 and other communicable diseases.

We found that use of any type of online grocery shopping (pick-up or delivery vs in-person) was similar regardless of rurality (rural, suburban, or urban), but that there was lower use of delivery and higher use of pickup among rural and suburban residents than urban residents. This may be a result of the geographical location of groceries stores that offer delivery wherein rural and suburban residents are outside of delivery areas but may still desire to shop online and go to the store for pickup. Also, many rural areas may only be served by small stores and farmers markets for which an online store may not be profitable or may lack the resources to create an online presence.

We hypothesized that SNAP participants would have lower use of online grocery shopping because of SNAP-specific barriers. SNAP participants can theoretically use their benefits online in all states except Alaska, Louisiana, and Montana.^14^ However, not all grocery stores are approved for SNAP online even if they accept SNAP at the brick and mortar location.^8, 14^ Also, SNAP benefits cannot be used to pay for delivery fees. Thus, for individuals without access to online payment methods (e.g., debit, credit cards), this prevents using online grocery shopping. Despite these barriers we found similar use of online grocery shopping among SNAP participants and non-participants. Our study was unable to explore SNAP participant specific barriers during the pandemic. However, there are still considerations to improve online grocery shopping that will increase the accessibility or affordability of online grocery shopping for SNAP participants. Policymakers could consider a universal platform for online shopping, which could help expand the number of stores that accept SNAP and participate in online grocery shopping. Furthermore, policymakers could allow the use of benefits to finance delivery fees and industry can set lower minimums to qualify for free delivery.

Lastly, there have been reports of shifting food habits and lifestyle during the pandemic, including increasing eating out of boredom, more snacking, and more energy dense foods as well as healthier eating.^15-19^ Our findings were similar to prior reports regarding more snacking and add to the literature that youth reported purchasing healthier foods more often than unhealthy foods, which may be related to the type of foods offered or promoted on online grocery shopping platforms.

Limitations of our study include that some youth may not be highly involved in grocery shopping. We framed the open-ended questions to focus on the youth and their families to minimize the influence of this limitation. Also, the cross-sectional nature of our study limits the ability to interpret causality for our findings. These changes in eating habits could be the result of many unmeasured factors including shifts in eating habits during the pandemic. Our study was not designed to study the etiology of these changes or measure these changes beyond the youths’ perceptions, thus these are interesting observations that require further inquiry. Lastly, the sample of SNAP participants and rural residents was small and we were unable to make further subgroup comparison (i.e. associations among rural SNAP participants compared to non-rural SNAP participants).

Our study provides timely insights into the use and perceived benefits and shortcomings of online grocery shopping during the COVID-19 pandemic from the perspective of youth. Our study found that online grocery shopping is feasible and accessible to many youth and their families, including those that may receive SNAP and live in rural areas. Healthcare providers and youth serving organizations can consider supporting youth and their families in using online grocery shopping to increase access to healthy foods and reduce the risk of transmission of COVID-19.

## Data Availability

The data are held within MyVoice (https://hearmyvoicenow.org/)

## Acknowledgements

NA

